# Does a History of Stroke Affect Outcomes in Non-Variceal Upper gastrointestinal Bleed?

**DOI:** 10.1101/2025.05.05.25327001

**Authors:** Anudeep Surendranath, Anupam K Gupta, Saurabh Singhal

## Abstract

**Background and Purpose:** Nonvariceal upper gastrointestinal bleeding (gastrointestinal) is potentially life-threatening. The study aimed to evaluate the clinical outcome in patients with upper gastrointestinal bleeding with a prior history of ischemic stroke.

**Methods:** The 2021 National Inpatient Sample database was employed to identify 259025 patients diagnosed with non-variceal upper gastrointestinal bleeding. 1485 patients exhibited a prior diagnosis of ischemic stroke. Data analysis was conducted using Stata version 18 to determine the primary outcome of mortality and secondary outcomes of length of hospitalization, cost, and post-hospitalization care needs.

**Results:** 259025 patients exhibited admissions due to non-variceal upper gastrointestinal bleed, and 1485 (0.57%) patients exhibited prior diagnoses of ischemic stroke. Patients with a history of ischemic stroke had a mean age was 72.35 years with higher comorbidities. Patients with a prior history of ischemic stroke had a higher risk of in-hospital mortality (OR 7.51,p<0.01). Length of hospitalization was longer by an mean of 5.68 days (p<0.01), and a higher need for discharge to a skilled nursing facility (OR 3.30, p<0.01). Race, median annual income, and geographical distribution were statistically noncontributory to the outcomes.

**Conclusion:** Patients with a history of ischemic stroke who present with non-variceal upper gastrointestinal bleeding tend to be older and have a higher comorbidity index. Prior history of stroke is an independent factor that contributes to the increased mortality in patients presenting with non-variceal upper gastrointestinal bleeding. They experience increased mortality rates, prolonged hospitalizations, higher costs during hospitalization, and a greater likelihood of being discharged to a nursing facility.

## Introduction

Stroke is a leading cause of mortality and significant morbidity in the United States and worldwide [1,2]. The lifetime risk of stroke is 25 percent [3]. Mortality and morbidity of stroke have improved because of early recognition and recanalization therapy with the use of intravenous or endovascular thrombolysis [4]. Multiple studies have shown efficacy in stroke management using Antithrombotic agents in short- and long-term management [5]. Aspirin alone or combined with clopidogrel/ oral anticoagulation is utilized for long-term management [5,6].

Upper gastrointestinal bleeding is bleeding in the gastrointestinal tract proximal to the ligament of Treitz and ranges from 48-160 per 100,000 adults with mortality of 3.3 to 4.3 per 10000 in the United States [7,8,9]. Upper gastrointestinal endoscopy is usually diagnostic and therapeutic[8]. A patient with upper gastrointestinal bleeding usually receives intravenous proton pump inhibitor therapy and blood transfusion. Patients on prior anticoagulation also undergo a reversal of anticoagulation if the bleeding is life-threatening [9,10,11].

gastrointestinal bleeding during stroke hospitalization is known to have poor outcomes and contributes to elevated mortality risk[12,13]. About 87% of all strokes are ischemic strokes[14]. It was well known that approximately 1.5% - 8 % of patients developed acute gastrointestinal bleed during admission for stroke. In a Japanese cohort, approximately 50% of cases had acute gastrointestinal bleed in the week after a stroke [12,13,14]. This is known to cause increased mortality and morbidity[13,14]. gastrointestinal bleeding has also been associated with an increased risk of stroke recurrence after a prior episode of acute ischemic stroke [14,15].

Risk factors for ischemic stroke, such as advanced age, male sex, black race, stress, hypertension, smoking, obesity, poor dietary habits and sedentary lifestyle, alcoholism, hyperlipidemia, associated medication, psychosocial factors, etc., can also predispose to the development of upper gastrointestinal bleeding [15,16].

During the management of upper gastrointestinal bleeding, multiple interventions, such as holding anticoagulation, reversal of anticoagulation, blood transfusion, and others, are physiologically and pharmacologically against long-term stroke prevention protocols that involve the use of antithrombotic agents for the management of ischemic stroke [11,16].

The purpose of the study was to identify outcomes in patients with gastrointestinal bleeding in patients with prior stroke.

## Methods

### Study design and database description

This study utilized the 2021 National Inpatient Sample (NIS), a comprehensive all-payer database developed by the Healthcare Cost and Utilization Project (HCUP) under the Agency for Healthcare Research and Quality (AHRQ). The NIS includes data from 48 participating states, covering 97% of the U.S. population and 96% of discharges from community hospitals, excluding rehabilitation and long-term acute care facilities. It is structured as a 20% stratified sample of hospital discharges, ensuring representativeness based on factors such as hospital ownership, bed capacity, teaching status, and geographic location. Its self-weighting design reduces error margins, enabling reliable national estimates while preserving patient confidentiality by excluding state and hospital identifiers. The substantial sample size enhances statistical power, facilitating the identification of differences in patient populations and outcomes, making it a valuable resource for healthcare research. As the data is de-identified and publicly available, Institutional Review Board (IRB) approval and informed consent were not required for this study.

### Study population

This study included all patients admitted with a principal diagnosis of non-variceal upper gastrointestinal (gastrointestinal) bleed, identified using the ICD-10 codes listed in the supplementary pages. The study population was divided into two groups based on the presence or absence of a history of ischemic stroke. We compared patient demographics and hospital characteristics between these groups to evaluate differences in clinical presentation and outcomes.

### Outcomes

The primary outcome of interest in this study was in-hospital mortality, comparing patients with and without a history of ischemic stroke who were admitted with a principal diagnosis of non-variceal upper gastrointestinal (gastrointestinal) bleeding. Secondary outcomes included total hospital length of stay (LOS), total hospital charges, and discharge disposition, which serve as critical metrics reflecting resource utilization, healthcare burden, and post-hospitalization care needs.

Statistical analysis: Data analysis was performed using Stata version 18. T-tests were used to calculate p-values for continuous variables, while Fisher’s exact test was applied to categorical and binary variables. To evaluate the association between a history of ischemic stroke and binary outcomes, multivariate logistic regression analysis was conducted, while multivariate linear regression was employed for continuous outcomes. Both models were adjusted for potential confounders to enhance the accuracy and reliability of the results.

## Results and Discussion

### Patient and hospital characteristics

The NIS dataset included 33.3 million observations, of which 259,025 were patients admitted with a principal diagnosis of non-variceal upper gastrointestinal (gastrointestinal) bleeding. Among these, 1,485 (0.57%) had a comorbid diagnosis of ischemic stroke. Table 1 offers a comprehensive comparison of patient and hospital characteristics for patients with and without an ischemic stroke diagnosis.

**Table 1:** Patient and hospital characteristics.

|  | No ischemic stroke | Ischemic stroke | P value |
| --- | --- | --- | --- |
| Female (%) | 120,000 (45.13) | 640 (43.1) | 0.49 |
| Male(%) | 139025 (53.67) | 845 (56.9) |  |
| Mean age (95%<br>conf. interval) | 66.55(66.35-66.75) | 72.35(71.00-73.69) | <0.01 |
| Race/ethnicity (%) |  |  | 0.07 |
| White | 170,000 (67.93) | 960 (67.37) |  |
| Black | 36,000 (14.53) | 240 (16.84) |  |
| Hispanic | 27,000 (10.82) | 100 (7.02) |  |
| Asian or Pacific<br>Islander | 8410 (3.35) | 70 (4.91) |  |
| Native American | 2675 (1.07) | 5 (0.35) |  |
| Other | 5795 (2.31) | 50 (3.51) |  |
| Median annual<br>income in patient's<br>zip code, US\$, no.<br>(%) | | | 0.21 |
| \$1 - \$51,999 | 78,000 (30.78) | 455 (30.95) | |
| \$52,000 - \$65,999 | 65,000 (25.64) | 345 (23.47) | |
| \$66,000 - \$87,999 | 60,000 (22.92) | 425 (28.91) | |
| \$88,000 or more | 50,000 (19.67) | 245 (16.67) | |
| Insurance type, no.<br>(%) |  |  | <0.01 |
| Medicare | 160,000 (63.56) | 1100 (76.12) |  |
| Medicaid | 36,000 (14.37) | 130 (9.0) |  |
| Private insurance,<br>including Health<br>Maintenance<br>Organizations<br>(HMO) | 44,000 (17.65) | 150 (10.38) |  |
| Self-pay | 11,000 (4.42) | 65 (4.5) |  |
| Hospital region, no.<br>(%) |  |  | 0.34 |
| Northeast | 42,000 (16.48) | 235 (15.82) |  |
| Midwest | 57,000 (22.17) | 395 (26.6) |  |
| South | 100,000 (39.51) | 560 (37.71) |  |
| West | 56,000 (21.84) | 295 (19.87) |  |
| Urban location (%) | 240,000 (92.87) | 1370 (92.26) | 0.67 |
| Teaching hospital | 190,000 (73.03) | 1120(75.42) | 0.36 |
| Charlson<br>Comorbidity Index<br>score, no. (%) |  |  | <0.01 |
| 0 | 2,000 (7.8) | 0 (0) |  |
| 1 | 56,000 (21.78) | 25 (1.68) |  |
| 2 | 45,000 (17.55) | 175 (11.78) |  |
| ≥3 | 140,000 (52.87) | 1285 (86.53) |  |

Patients with a history of ischemic stroke were significantly older, with a mean age was 72.35 years (95% CI: 71.00–73.69) compared to 66.55 years (95% CI: 66.35–66.75) (p<0.01). The proportion of female patients was similar between the groups (43.1% vs. 45.13%, p=0.49).

Racial distribution did not significantly differ (p=0.07), with White patients comprising the majority (67.37% stroke vs. 67.93% no stroke). Median household income distribution also did not differ significantly (p=0.21). However, insurance status varied significantly (p<0.01), with Medicare coverage being more common among patients with a history of ischemic stroke (76.12% vs. 63.56%), while Medicaid (9.0% vs. 14.37%) and private insurance (10.38% vs. 17.65%) were less frequent.

Hospital characteristics, including region (p=0.34), urban location (p=0.67), and teaching hospital status (p=0.36), were similar between groups. However, patients with a history of ischemic stroke had a significantly elevated Charlson Comorbidity Index score (p<0.01), with 86.53% having a score ≥3 compared to 52.87% in the non-stroke group, indicating a greater overall burden of comorbidities.

These findings indicate that patients with a history of ischemic stroke admitted with non-variceal gastrointestinal bleeding are older, have a a more substantial burden of comorbid conditions burden, and are more likely to be covered by Medicare, which may influence clinical management and outcomes.

### Primary outcome

Among the 259,025 patients admitted with a principal diagnosis of non-variceal upper gastrointestinal (gastrointestinal) bleeding, 5,430 (2.1%) died during hospitalization. Table 2 offers a comprehensive comparison of in-hospital mortality between patients with and without a history of ischemic stroke.

**Table 2:** Primary outcome.

| Outcome | Exposure | Odds Ratio (OR) | 95% Confidence Interval (CI) | p-value |
| --- | --- | --- | --- | --- |
| Mortality (Unadjusted) | Ischemic stroke | 7.51 | 5.42-10.39 | <0.01 |
| Mortality (Adjusted) | Ischemic stroke | 4.96 | 3.47-7.09 | <0.01 |

Patients with a history of ischemic stroke had a significantly elevated risk of in-hospital mortality, with an unadjusted odds ratio (OR) of 7.51 (95% CI: 5.42–10.39, p<0.01). After adjusting for age, sex, race, median household income, insurance type, hospital region, urban versus rural location, teaching hospital status, and Charlson Comorbidity Index score, the association remained statistically significant, with an adjusted OR of 4.96 (95% CI: 3.47– 7.09, p<0.01).

These results indicate that a history of ischemic stroke is associated with increased in-hospital mortality among patients admitted with non-variceal gastrointestinal bleeding, as detailed in Table 2.

### Secondary outcomes

The mean length of stay for patients admitted with a principal diagnosis of non-variceal gastrointestinal bleed was 4.67 days. After adjusting for confounders, patients with ischemic stroke had an average hospital stay that was 5.68 days longer than those without a history of stroke (95% CI: 4.61–6.76; p<0.01).

The average total hospital charges for patients admitted with non-variceal gastrointestinal bleed were $61,836. After adjustment, hospital charges for patients with ischemic stroke were $100,211.7 higher compared to those without ischemic stroke (95% CI: $77,435.08– $122,988.3; p<0.01).

Regarding discharge disposition, patients with ischemic stroke had significantly lower odds of discharge home (OR: 0.23, 95% CI: 0.18–0.30; p<0.01) and increased likelihood of discharge to a skilled nursing facility (OR: 3.30, 95% CI: 2.54–4.30; p<0.01).

Table 3 presents a detailed comparison of secondary outcomes, including hospital length of stay, total hospital charges, and discharge disposition among patients with and without a history of ischemic stroke.

**Table 3:**
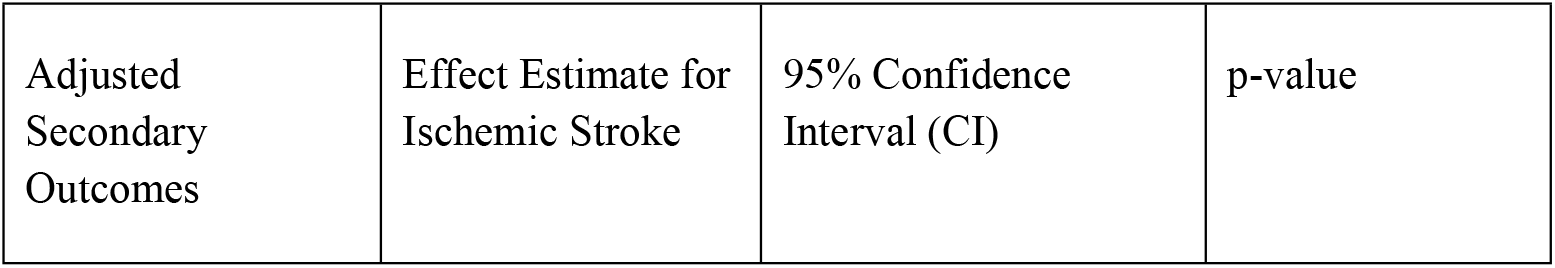
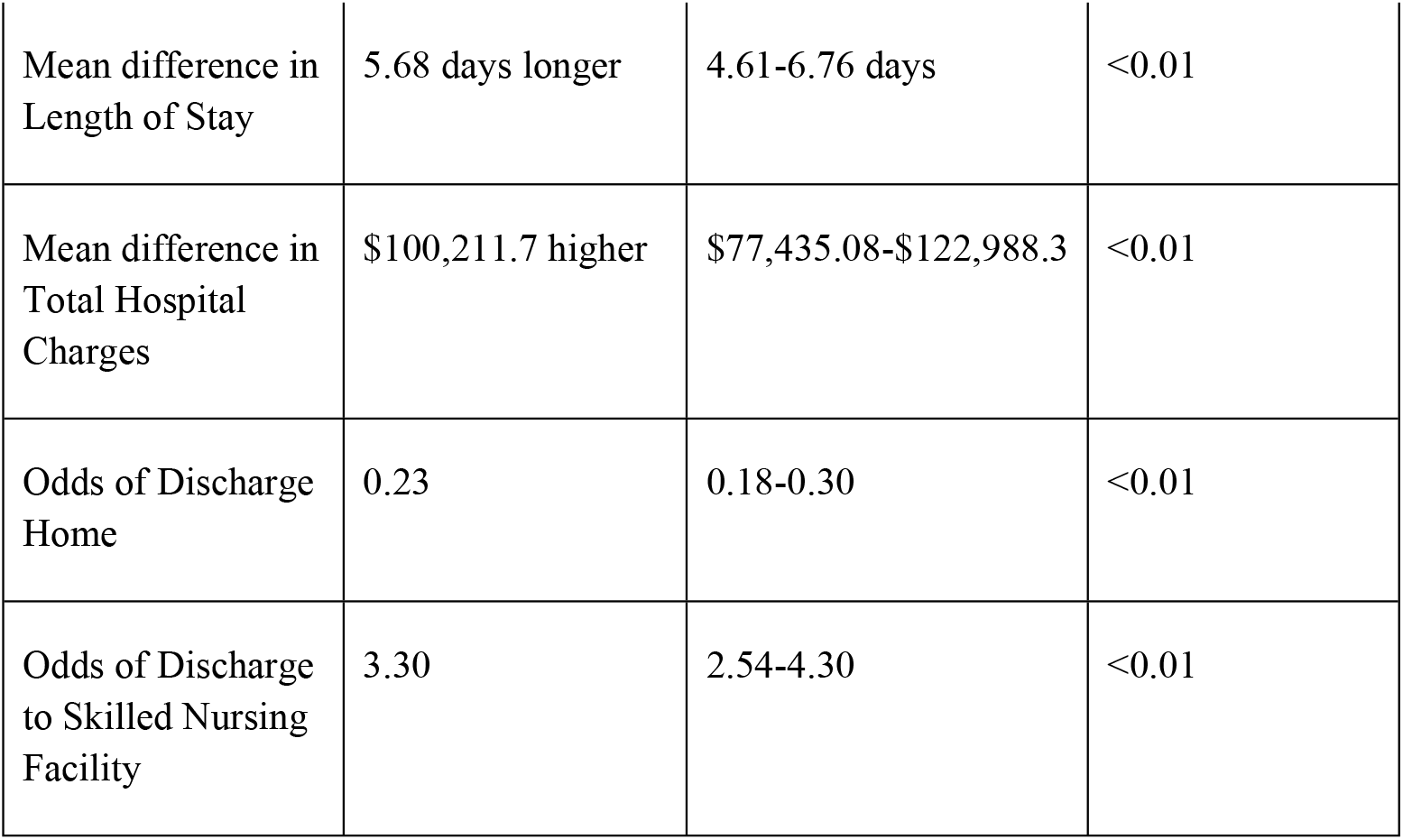
Secondary outcomes.

| Adjusted Secondary Outcomes | Effect Estimate for Ischemic Stroke | 95% Confidence Interval (CI) | p-value |
| --- | --- | --- | --- |
| Mean difference in Length of Stay | 5.68 days longer | 4.61-6.76 days | <0.01 |
| Mean difference in Total Hospital Charges | \$100,211.7 higher | \$77,435.08-\$122,988.3 | <0.01 |
| Odds of Discharge Home | 0.23 | 0.18-0.30 | <0.01 |
| Odds of Discharge to Skilled Nursing Facility | 3.30 | 2.54-4.30 | <0.01 |

## Discussion

The NIH database has 33.3 million observations. As per the 2021 database, there were 259025 patients admitted to the hospital due to a primary non-variceal upper gastrointestinal bleed. Only 0.57% of patients exhibited a prior history of ischemic stroke. These patients with prior history of stroke and presenting with gastrointestinal bleed had a higher mean age was 72.35 years versus patients presenting with acute upper gastrointestinal bleed, and this correlation was significant. These patients also had a more substantial burden of comorbid conditions at the time of presentation, which was statistically significant.

Race, median household income, geographical location, and local hospital availability were not statistically relevant in the statistical analysis.

Acute upper gastrointestinal bleeding is a serious complication in patients with a prior history of stroke, and most studies have estimated it between 1-5% [18,19]. However, our database was below 1%.

Patients in this cohort of gastrointestinal bleeding with prior strokes are usually older, with a more substantial burden of comorbid conditions and with alteration in level of consciousness [20,21,22]. Many of the patients exhibited had a prior history of gastrointestinal bleeding with the use of medications like NSAIDs and anticoagulants, which predispose them to recurrent episodes of bleeding [23]. Having a more substantial burden of comorbid conditions predisposes them to have more medications, changes in the level of consciousness, H pylori infection, and neurological factors causing predisposition to stress ulcers [24,25,26]. Many of these patients also have a prior history of atrial fibrillation, which is also an independent factor in predisposition to gastrointestinal bleeding [27].

Patients with non-variceal upper gastrointestinal bleeding with a prior history of stroke have a higher risk of in-hospital mortality, which could be attributed to advanced age, comorbidities, mental state, etc [28,29].

We adjusted our statistical analysis for age, sex, median household income, insurance type, hospital location (urban vs. local), and comorbidity. The results remained statistically significant with an odds ratio of 4.96 (95% CI: 3.47–7.09, p<0.01). These findings indicate that patients with a prior history of stroke presenting with non-variceal gastrointestinal bleeding have elevated mortality risk, with stroke being an independent risk factor for outcome.

Secondary outcomes show patients exhibited more extended hospital stays, hospital charges, and higher medical needs at the time of discharge. These analyses were statistically significant. Patients with an upper gastrointestinal bleed and a prior history of stroke are a small fraction of affected individuals; however, they tend to be older, sicker, and more complicated due to these factors.

Patients with a prior history of strokes, depending on the severity, comorbidity, and mental status, require a higher level of attention and care in the management.

## Conclusion

Among patients admitted with a primary diagnosis of acute non-variceal upper gastrointestinal bleeding, 0.57% had a prior history of ischemic stroke. These patients exhibited a mean age was over 72 years. They experienced higher in-hospital mortality and morbidity, with an adjusted odds ratio of 4.96 (p < 0.01). Prior history of stroke is an independent factor in the mortality of patients presenting with non-variceal upper gastrointestinal bleed.

Additionally, they typically had a longer hospitalization duration, averaging 5.68 days longer (p < 0.01), resulting in elevated healthcare expenditures estimated at approximately $10000. At discharge, they also had a greater dependence on skilled nursing facilities.

## Limitations

This study has several limitations. First, as it relies on data from the National Inpatient Sample, which consists of administrative discharge-level records, it assumes accurate coding of diagnoses and procedures. However, the potential for misclassification of diagnoses cannot be entirely ruled out. Although the NIS represents approximately 20% of all hospital discharges and employs stratified sampling with weighting to generate national estimates, it remains a sample rather than a complete census. As a result, certain nuanced patient- or hospital-level characteristics may not be fully captured.

Additionally, the NIS does not provide clinical data, such as timeline, temporal association, medication use, laboratory test results, etc. Lastly, the retrospective nature of this study restricts our ability to establish causal relationships between a history of ischemic stroke and the observed outcomes. Future research incorporating prospective study designs and additional clinical variables would be beneficial in further elucidating these associations

## Data Availability

https://hcup-us.ahrq.gov/tools_software.jsp
The data used in this study is publicly available, de-identified, and contains no personal identifiers. As such, no ethics committee or IRB approval was required for its use in research.

https://hcup-us.ahrq.gov/tools_software.jsp

